# Simple Laboratory and Clinical Parameters as Predictor for Electrocardiographic Abnormalities Among Hospitalized Patients with Chronic Kidney Disease: A Cross-Sectional Study

**DOI:** 10.1101/2021.01.15.21249916

**Authors:** Rerdin Julario, Ricardo Adrian Nugraha, Bagus Putra Dharma Khrisna, Tony Santoso Putra, Eka Prasetya Budi Mulia, Ryan Enast Intan, Firas Farisi Alkaff

## Abstract

**Background:** In developing countries, even electrocardiography (ECG) hasn’t been used widely in most health-care centers. The ability of physicians to refer to chronic kidney disease (CKD) patients for ECG, often collide with several barriers and costs. Therefore, we need to formulate the simplest and most efficient model to predict when CKD patients need to be referred due to potential ECG abnormalities.

**Objective:** The aim of this study was to develop several clinical and laboratory parameters as a predictor of any ECG abnormalities.

**Materials and Methods:** A retrospective cross-sectional study design held at Dr. Soetomo General Academic Hospital, Surabaya, Indonesia. Subjects were hospitalized patients with CKD between 1 January to 31 December 2019. 198 CKD patients (101 males) were enrolled for the study. All patients had demographic information, detailed clinical profile, resting 12-lead ECG recording, complete blood count, serum electrolyte and renal function test profile during admission and results were interpreted blindly by two cardiologists. Statistical analysis was done by SPSS 17.0.

**Results:** A total of 198 patients were included in this study. Mean ages were 52.2±11.8 years old and fifty-one percent were males. Eighty-eight percent of patients from 198 patients had ECG abnormality. AUC of hemoglobin level to discriminate poor R wave progression, pathological Q wave, non-spesific ST-T changes, and frontal axis deviation were 0.532, 0.641, 0.556 and 0.693, respectively. In multivariate logistic regression analysis, only higher systolic blood pressure was determined as an independent predictor of abnormal ECG finding in CKD patients, as systolic blood pressure increase by one unit, the odds of having abnormal ECG is increased 1.02 times (95% CI: 1.00 – 1.02, *p*=0.042).

**Conclusion:** The ECG abnormalities can be found in hospitalized CKD patients. Fragmented QRS and long QTc were the highest prevalent ECG abnormalities in our study. Serum creatinine and hemoglobin could predict peaked T wave and prolonged QTc among hospitalized CKD patients. Systolic blood pressure could predict prolonged QTc and fragmented QRS in CKD patients.

## INTRODUCTION

Chronic kidney disease (CKD) is associated with various kinds of complications that lead to poor health outcomes. The risk of cardiovascular disease (CVD) has increased in patients with CKD even before reaching End-Stage Renal Disease (ESRD). The mortality rate in dialysis patients due to cardiovascular complications is high. The prognosis in developing countries is poor due to inadequate diagnostic and late presentation[1].

KDIGO 2012 recommends active intervention by paying attention to the conditions of CVD and related comorbidities to reduce hospitalization and mortality for people with CKD[2]. Based on the guideline principles, CKD patients need to be closely monitored for early signs and symptoms of CVD. Even annual ECG monitorings are mandatory to detect any sign of CVD at an early stage. Unfortunately, many clinicians, particularly in developing countries, face a big problem to adopt these recommendations [3]. Such as electrocardiography (ECG) hasn’t been used widely in most health-care centers. Therefore, patients need to be referred to bigger health-care centers or hospitals. On the other hand, the ability of physicians to refer chronic kidney disease (CKD) patients for ECG examination or comprehensive specialist treatment, often collide with the geographical barrier, accessibility, patients’ belief, policy, cost, and insurance[3]. Patients with CKD in developing countries usually seek medical advice after falling at an advanced stage due to a lack of awareness, education, financial constraints, and medical facilities[4, 5].

Due to several barriers, the decision of whether to refer CKD patients to comprehensive specialist treatment or even for ECG measurement is challenging and confusing for many clinicians. In addition, only a few studies provide information on the frequency of ECG changes in the local CKD population which can make the physician less aware of the complications of CVD in CKD[6]. The frequency and predictor of ECG abnormalities in CKD patients in Indonesia may also be different from the literature that has been reported[7]. Therefore, we need to formulate the simplest and most efficient model from the clinical and laboratory parameters to predict the time when CKD patients need to be referred due to potential ECG abnormalities. Since there is a lack of Indonesian-based research about predictor model for ECG abnormalities on CKD patients, we develop our model from several clinical and simple laboratory parameters to predict any ECG abnormalities in CKD patients treated in a referral hospital in Indonesia.

## MATERIALS AND METHODS

### Ethical clearance

Institutional ethics and research committee approved the study. Dr. Soetomo General Academic Hospital conferred ethical clearance for this study. Compliance with ethical standards, this research proposal was approved by the ethical committees of Dr. Soetomo General Academic Hospital in collaboration with Universitas Airlangga College of Medicine Research Ethics Council (Ref: 1811/KEPK/II/2020) under the name of Rerdin Julario as the Principal Investigator.

### Study design and study setting

This study was an analytic observational study using a retrospective cross-sectional study design. This study held at Dr. Soetomo General Academic Hospital, Surabaya, Indonesia. Dr. Soetomo General Academic Hospital is the biggest hospital in East Java and also the referral hospitals for East Indonesia region with an academic affiliation with Faculty of Medicine, Universitas Airlangga. All patients between the ages of 15–90 years with known or newly diagnosed CKD admitted at the internal medicine ward were included. The study was conducted over 12 months period from 1 January to 31 December 2019.

### Population and sample

All patients in different stages of CKD were included in this study. Patients with unstable hemodynamic, history of cardiac arrest, previously consumed antiarrhythmic drugs and lack of complete medical record data were excluded. This study used consecutive sampling.

### Outcomes and variables

Potential cases of CKD were identified by an eGFR 60 ml/min per 1.73 m^2^. To predict the risk of CV in previous epidemiological studies, we can use an eGFR threshold of 60 ml / min per 1.73 m^2^ [2]. Estimated GFR (eGFR) was calculated using the CKD-EPI equation [8]. Medical records, Patient’s history, and laboratory information were reviewed to gain data on patient’s age, gender, history of hyperglycemia, hypertension, resting heart rate, systolic and diastolic blood pressure, serum creatinine, blood ureum nitrogen, eGFR, electrolytes, and complete blood count. The patient was classified to have ECG abnormalities if one of the ECG parameters didn’t fulfill the reference range.

### Data collection

All patients had resting 12-lead ECG examination. All ECGs were reviewed with the following parameters: heart rhythm, heart rate, frontal axis, P wave, PR interval, QRS duration, pathological Q wave, fragmented QRS morphology, QTc interval, ST-segment deviation, and T wave morphology; by two independent consultant physicians. The aforementioned parameters were studied and measured manually and compared with published normal values. The normal limit and definition used for this study were shown in table 1 and table 2. The abnormal findings were defined if were away from minimal or maximal limits or normal morphology. Precordial lead subjective assessments of the peaking of the T wave were also noted. Subsequent ECGs if done were not analyzed for study purposes. Descriptive results were presented in frequency and percentage. Analytic results were presented in ROC curve.

**Table 1.**
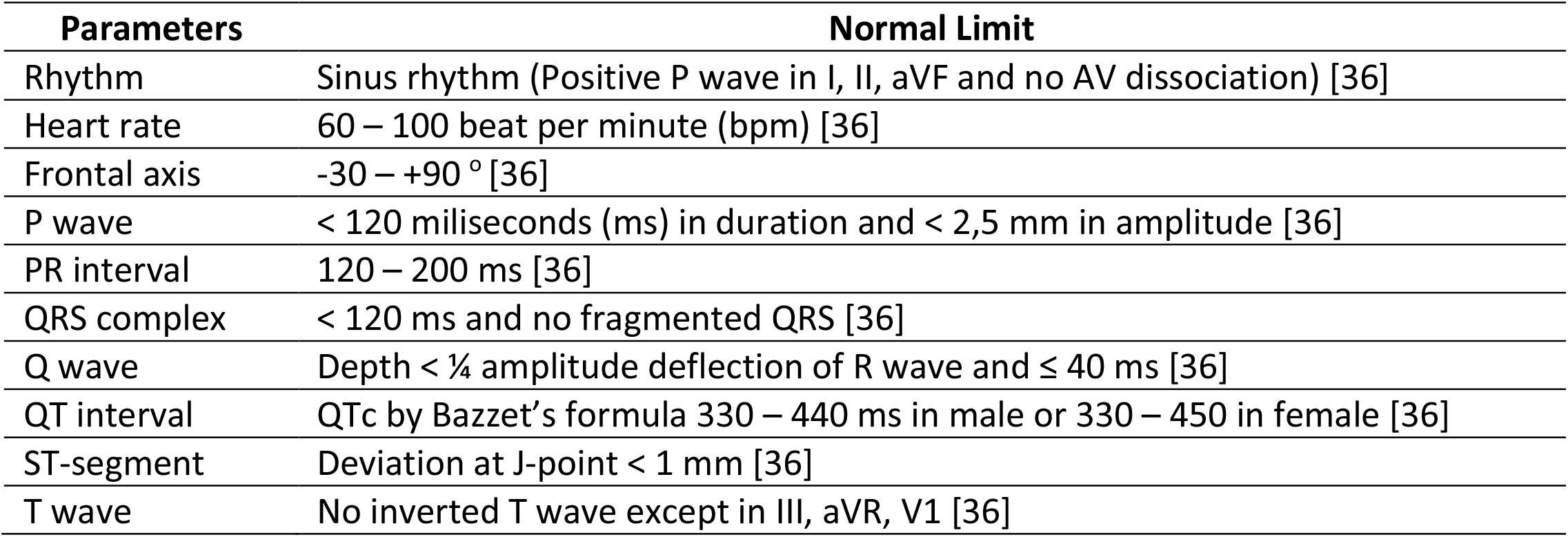
Normal limit of ECG parameters

**Table 2.**
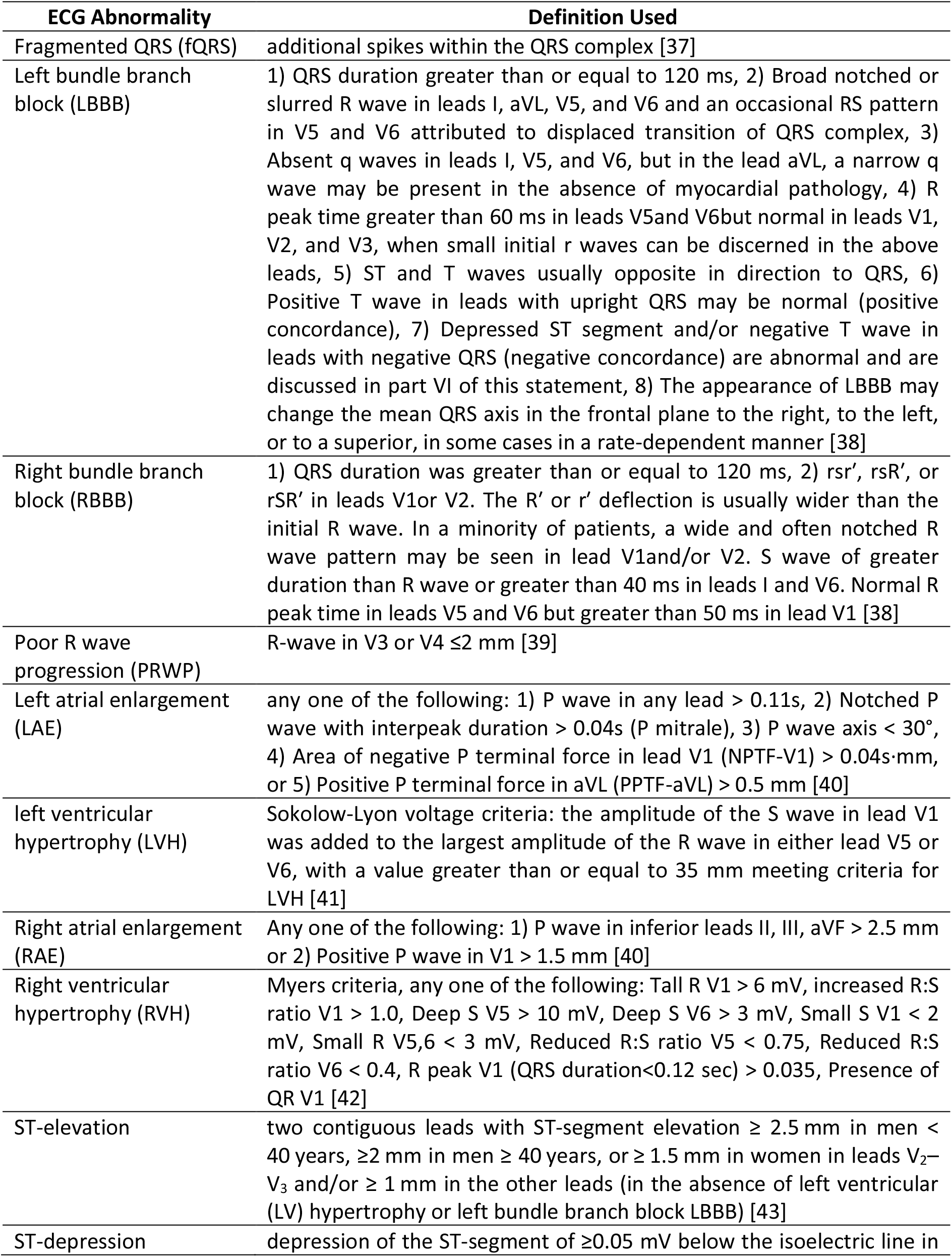
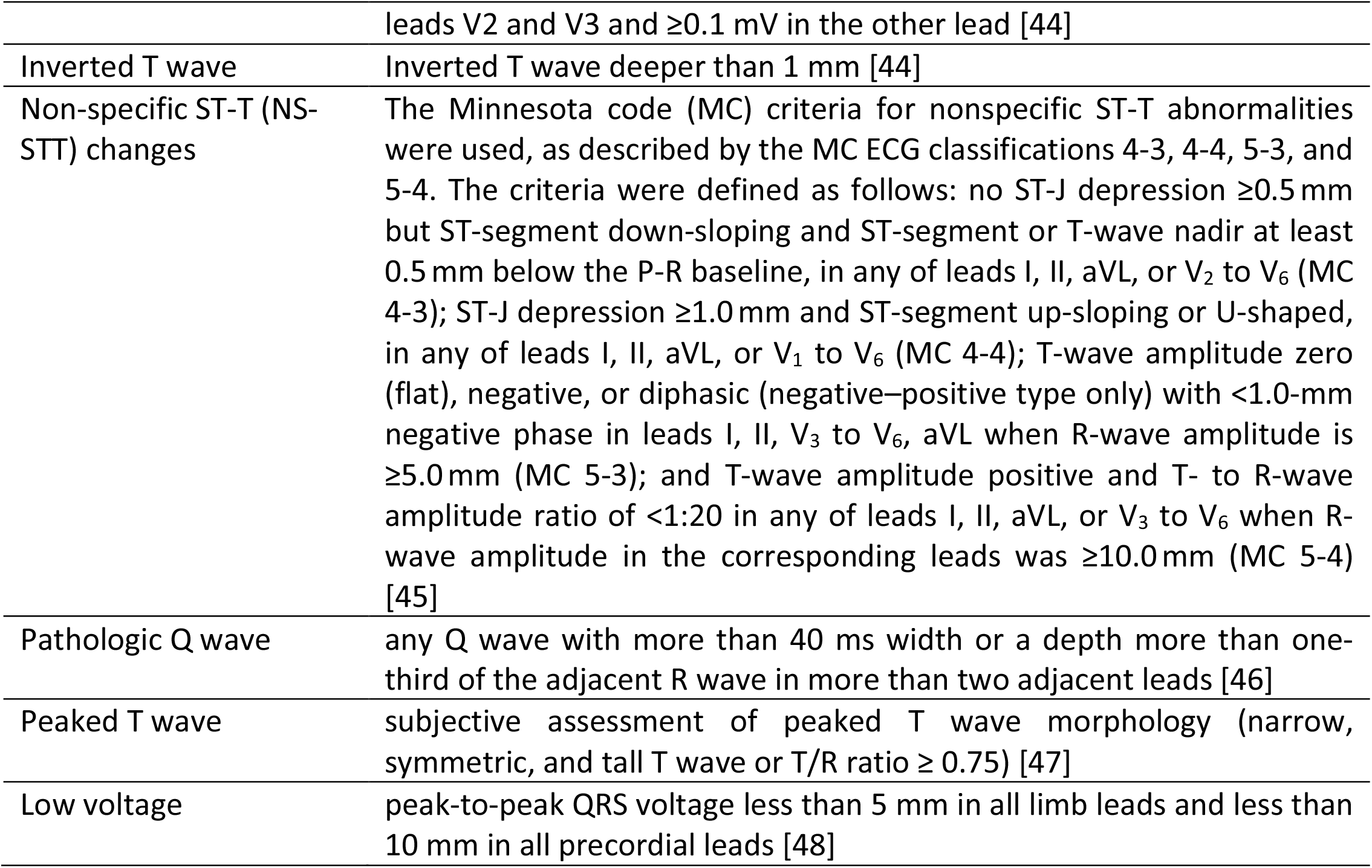
Definition used for ECG abnormality

### Statistical analysis

For bivariate analysis, χ^2^-test was used for analyzing categorical data, and independent t-test or Mann-Whitney U test was used for continuous data as appropriate. For univariate and multivariate analysis, the logistic regression model was performed with backward selection. Model calibration was tested with the Hosmer-Lemeshow test. The accuracy of the test was evaluated based on ROC curve analysis. The results on the level of *p* < 0.05 were assumed to be statistically significant. All statistical analyses were performed using SPSS 26.0 (IBM SPSS Statistics for Windows, IBM Corporation, Armonk, NY).

## RESULTS

### Baseline characteristic of the study

A total of 198 patients were included in this study. Characteristics of the study population are summarized in (Table 3). The mean age of all patients was 52.2±11.8 years old. A total of 101 patients (51%) were males. Mean serum creatinine was 10.5±8.0 mg/dL and mean eGFR was 10.6±14.4 mL/min/1.73 m^2^. Most patients were in stage 5 of CKD, which was 164 patients (82.8%). 56.1% (111 patients from 198 patient) patients had hypertension, 37.9% had diabetes mellitus, and 24.7% had a known history of CVD. The mean systolic and diastolic blood pressure was 142.1±28.2 mmHg and 83.7±16.3 mmHg respectively. 143 (72.2%) had anemia with mean hemoglobin level was 8.1±2.3 g/dL. As the result, the ECG abnormality can be found in 176 or almost 88.9%. Another abundant comorbidity was metabolic acidosis (31.8%). Most patients had high (45.5%) or normal (44.9%) serum potassium, and only 9.1% of patients had low serum potassium, with mean serum potassium was 4.8±1.06 mEq/L.

**Table 3.** Baseline characteristic of study population

| **Variables** | **N (198)** | **Proportion (%)** |
| --- | --- | --- |
| **Age Range** |  |  |
| <= 30 | 10 | 5.1% |
| 31 - 40 | 25 | 12.6% |
| 41 - 50 | 39 | 19.7% |
| 51 - 60 | 79 | 39.9% |
| 61 - 70 | 35 | 17.7% |
| > 70 | 10 | 5.1% |
| **Sex** |  |  |
| Male | 101 | 51% |
| Female | 97 | 49% |
| **Comorbidities** |  |  |
| Anemia | 143 | 72.2% |
| HTN | 111 | 56.1% |
| Diabetes Mellitus | 75 | 37.9% |
| Metabolic Acidosis | 63 | 31.8% |
| Pneumonia | 48 | 24.2% |
| Hypoalbuminemia | 48 | 24.2% |
| Hyponatremia | 38 | 19.2% |
| Pleural Effusion | 30 | 15.2% |
| Pulmonary Edema | 29 | 14.7% |
| CVD, others | 16 | 8.1% |
| Nephrolithiasis | 14 | 7.1% |
| Heart Failure | 6 | 3.0% |
| Coronary Artery Disease | 1 | 0.5% |
| Pericardial Effusion | 1 | 0.5% |
| **GFR mL/min/1.73 m<sup>2</sup> (CKD stage)** |  |  |
| > 89 (stage 1) | 1 | 0.5% |
| 60-89 (stage 2) | 2 | 1% |
| 45-59 (stage 3A) | 2 | 1% |
| 30-44 (stage 3B) | 9 | 4.6% |
| 15-29 (stage 4) | 23 | 11.7% |
| <15 (stage 5) | 160 | 81.22% |
| **Potassium level (mEq/L)** |  |  |
| <3.5 | 18 | 9.1% |
| 3.5-5.0 | 89 | 44.9% |
| >5.0 | 90 | 45.5% |

### Abnormal ECG findings

Various ECG changes are observed in Table 4

**Table 4.** ECG Abnormalities

| **Abnormalities** | **N (Proportion)** |
| --- | --- |
| Long QTc | 75 (37.9%) |
| Fragmented QRS complex | 59 (29.8%) |
| Poor Precordial R wave Progression | 48 (24.2%) |
| Left Atrial Enlargement | 46 (23.2%) |
| Peaked T wave | 43 (21.7%) |
| Left Ventricular Hypertrophy | 31 (15.6%) |
| Pathological Q wave | 27 (13.6%) |
| Non-specific ST-T Changes | 26 (13.1%) |
| Frontal Axis Deviation | 21 (10.6%) |
| Inverted T Wave | 14 (7.1%) |
| 1<sup>st</sup> Degree AV Block | 14 (7.1%) |
| ST Segment Depression | 13 (6.6%) |
| Right Bundle Branch Block | 11 (5.6%) |
| Wide QRS Complex | 7 (3.5%) |
| Premature Ventricular Contraction (PVC) | 6 (3.0%) |
| Right Ventricular Hypertrophy | 5 (2.5%) |
| Non-Sinus Rhythm | 4 (2.0%) |
| Low voltage | 3 (1.5%) |
| ST segment elevation | 2 (1.0%) |
| Right Atrial Enlargement | 2 (1.0%) |
| Premature Atrial Contraction (PAC) | 1 (0.5%) |
| Short QTc | 1 (0.5%) |
| Left Bundle Branch Block | 1 (0.5%) |
| 2<sup>nd</sup> Degree AV Block | 1 (0.5%) |

### Cut-off value for serum creatinine and other biomarkers to discriminate long QTc interval

The cut-off value of serum creatinine to discriminate long QTc can be seen in ROC Curve in figure 1. For bivariate analysis, serum creatinine is positively discriminate long QTc based on ROC curve (Figure 1) with area under curve (AUC) 0.544 (*P*<0.001) with the best cut off serum creatinine level at 7.58 (sensitivity 62% and specificity 50%) (Table 5).

**Table 5.** Area under the curve (AUC) of clinical prediction model for age, BUN, serum creatinine, eGFR, systolic blood pressure, diastolic blood pressure, hemoglobin level and serum potassium. Prediction of long QT interval by age, BUN, serum creatinine, eGFR, systolic blood pressure, diastolic blood pressure, hemoglobin level and serum potassium. *P* < 0.001 for all ROC curves compared with 0.5 curves.

| Test Result Variable(s) | Area Under the Curve |  |  |  |  |
| --- | --- | --- | --- | --- | --- |
|  | Area | Std. Error<sup>a</sup> | Asymptotic Sig.<sup>b</sup> | Asymptotic 95% Confidence Interval |  |
|  |  |  |  | Lower Bound | Upper Bound |
| Age | ,518 | ,042 | ,672 | ,436 | ,600 |
| BUN | ,395 | ,040 | ,013 | ,316 | ,475 |
| Creatinin | ,554 | ,042 | ,199 | ,472 | ,636 |
| eGFR | ,450 | ,042 | ,232 | ,368 | ,532 |
| Systole | ,555 | ,041 | ,191 | ,474 | ,636 |
| Diastole | ,540 | ,041 | ,343 | ,459 | ,621 |
| Hemoglobin | ,511 | ,042 | ,799 | ,429 | ,592 |
| Kalium | ,379 | ,042 | ,004 | ,296 | ,462 |

The test result variable(s): Age, BUN, Creatinin, Systole, Diastole, Hemoglobin, Kalium has at least one tie between the positive actual state group and the negative actual state group. Statistics may be biased.
a. Under the nonparametric assumption
b. Null hypothesis: true area = 0.5

**Figure 1.**
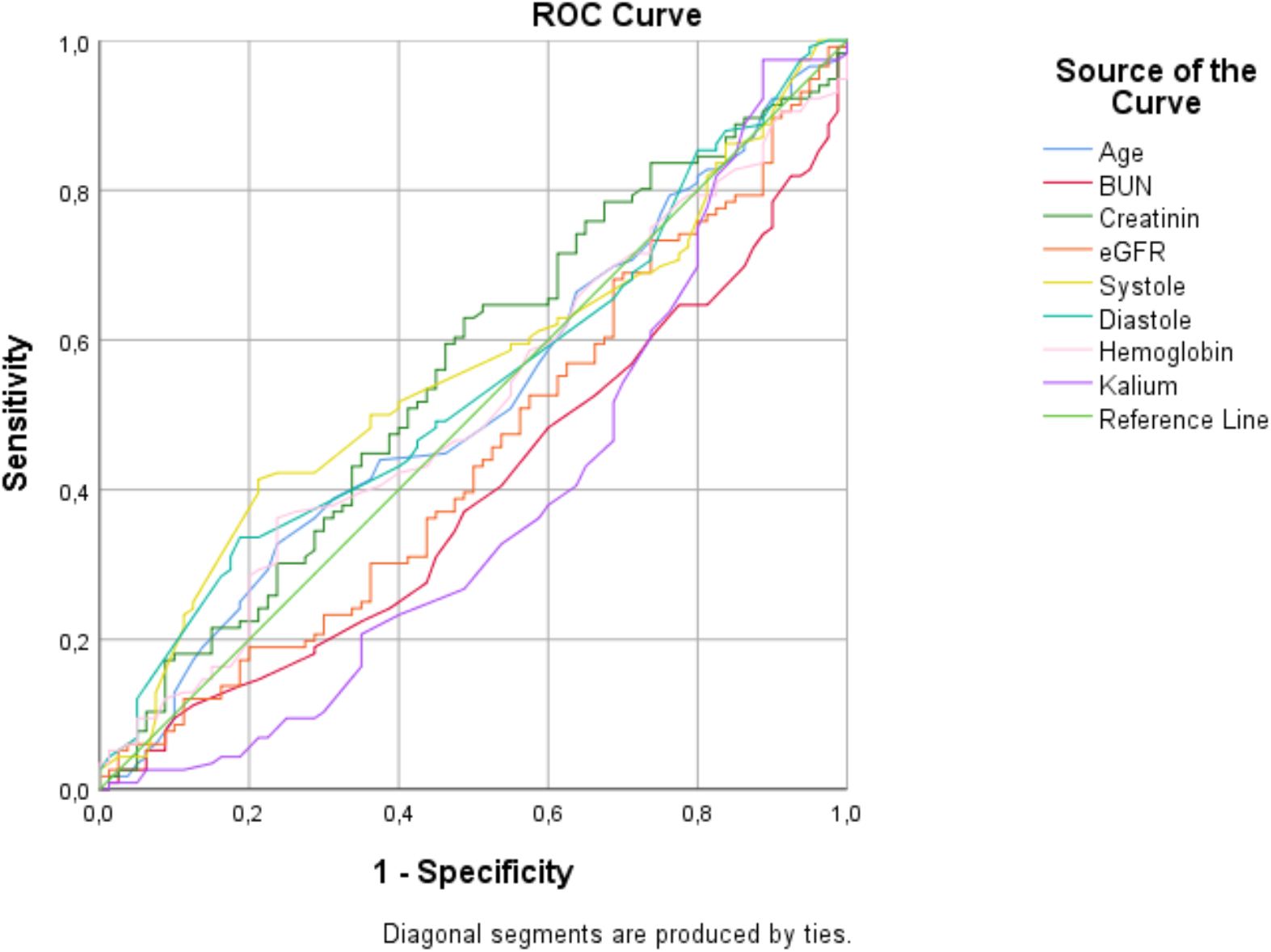
Receiver operator characteristic analysis of the clinical prediction model for age, BUN, serum creatinine, eGFR, systolic blood pressure, diastolic blood pressure, hemoglobin level and serum potassium. Prediction of long QT interval by age, BUN, serum creatinine, eGFR, systolic blood pressure, diastolic blood pressure, hemoglobin level and serum potassium. *P* < 0.001 for all ROC curves compared with 0.5 curves.

### Cut-off value for serum creatinine and other biomarkers to discriminate fragmented QRS

The cut-off value of serum creatinine to discriminate fragmented QRS can be seen in ROC Curve (Figure 2). For bivariate analysis, serum creatinine is a negative predictor for fragmented QRS based on ROC curve with area under curve (AUC) 0.496 (Table 6).

**Table 6.** Area under the curve (AUC) of clinical prediction model for age, BUN, serum creatinine, eGFR, systolic blood pressure, diastolic blood pressure, hemoglobin level and serum potassium. Prediction of fragmented QRS by age, BUN, serum creatinine, eGFR, systolic blood pressure, diastolic blood pressure, hemoglobin level and serum potassium. *P* < 0.001 for all ROC curves compared with 0.5 curves.

| Test Result Variable(s) | Area Under the Curve |  |  |  |  |
| --- | --- | --- | --- | --- | --- |
|  | Area | Std. Error<sup>a</sup> | Asymptotic Sig.<sup>b</sup> | Asymptotic 95% Confidence Interval |  |
|  |  |  |  | Lower Bound | Upper Bound |
| Age | ,502 | ,045 | ,964 | ,413 | ,591 |
| BUN | ,473 | ,046 | ,553 | ,384 | ,563 |
| Creatinin | ,500 | ,046 | 1,000 | ,410 | ,590 |
| eGFR | ,502 | ,046 | ,969 | ,411 | ,592 |
| Systole | ,532 | ,048 | ,478 | ,439 | ,626 |
| Diastole | ,476 | ,048 | ,589 | ,381 | ,570 |
| Hemoglobin | ,545 | ,046 | ,321 | ,455 | ,635 |
| Kalium | ,496 | ,047 | ,926 | ,405 | ,587 |

The test result variable(s): Age, BUN, Creatinin, Systole, Diastole, Hemoglobin, Kalium has at least one tie between the positive actual state group and the negative actual state group. Statistics may be biased.
a. Under the nonparametric assumption
b. Null hypothesis: true area = 0.5

**Figure 2.**
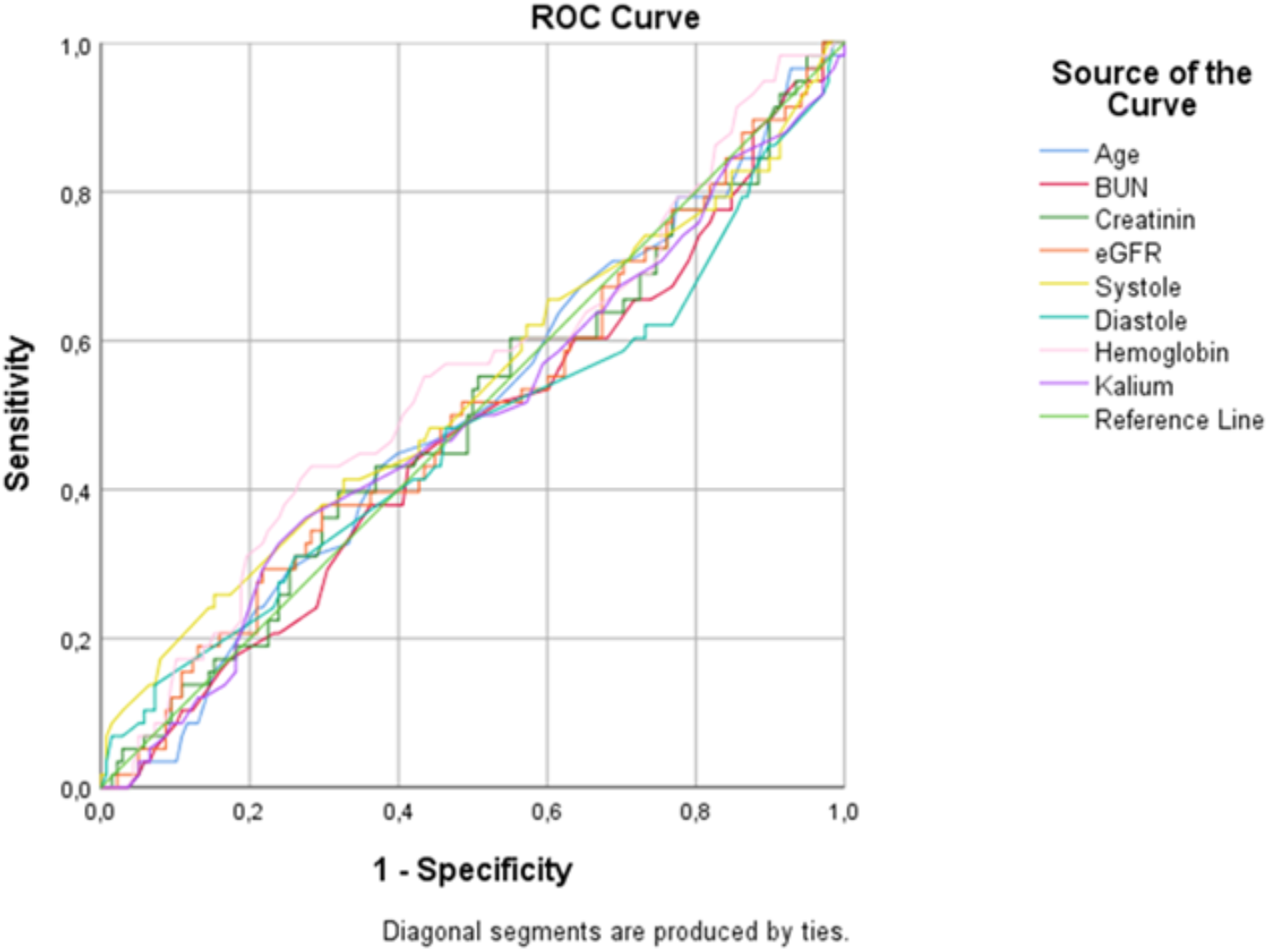
Receiver operator characteristic analysis of the clinical prediction model for age, BUN, serum creatinine, eGFR, systolic blood pressure, diastolic blood pressure, hemoglobin level and serum potassium. Prediction of fragmented QRS by age, BUN, serum creatinine, eGFR, systolic blood pressure, diastolic blood pressure, hemoglobin level and serum potassium. *P* < 0.001 for all ROC curves compared with 0.5 curves.

### Cut-off value for serum creatinine and other biomarkers to discriminate left ventricle hypertrophy

The cut-off value of serum creatinine to discriminate left ventricle hypertrophy (LVH) can be seen in ROC Curve (Figure 3). For bivariate analysis, serum creatinine is positively discriminate LVH based on ROC curve with area under curve (AUC) 0.635 (*P*<0.001) with the best cut off serum creatinine level at 7.045 (sensitivity 81% and specificity 46%) (Table 7).

**Table 7.** Area under the curve (AUC) of clinical prediction model for age, BUN, serum creatinine, eGFR, systolic blood pressure, diastolic blood pressure, hemoglobin level and serum potassium. Prediction of LVH by age, BUN, serum creatinine, eGFR, systolic blood pressure, diastolic blood pressure, hemoglobin level and serum potassium. *P* < 0.001 for all ROC curves compared with 0.5 curves.

| Test Result Variable(s) | Area Under the Curve |  |  |  |  |
| --- | --- | --- | --- | --- | --- |
|  | Area | Std. Error<sup>a</sup> | Asymptotic Sig.<sup>b</sup> | Asymptotic 95% Confidence Interval |  |
|  |  |  |  | Lower Bound | Upper Bound |
| Age | ,519 | ,061 | ,733 | ,399 | ,639 |
| BUN | ,402 | ,057 | ,080 | ,290 | ,514 |
| Creatinin | ,634 | ,048 | ,017 | ,540 | ,728 |
| eGFR | ,388 | ,050 | ,046 | ,291 | ,486 |
| Systole | ,578 | ,052 | ,162 | ,475 | ,681 |
| Diastole | ,622 | ,050 | ,029 | ,524 | ,720 |
| Hemoglobin | ,447 | ,052 | ,347 | ,345 | ,550 |
| Kalium | ,467 | ,055 | ,551 | ,359 | ,574 |

The test result variable(s): Age, BUN, Creatinin, Systole, Diastole, Hemoglobin, Kalium has at least one tie between the positive actual state group and the negative actual state group. Statistics may be biased.
a. Under the nonparametric assumption
b. Null hypothesis: true area = 0.5

**Figure 3.**
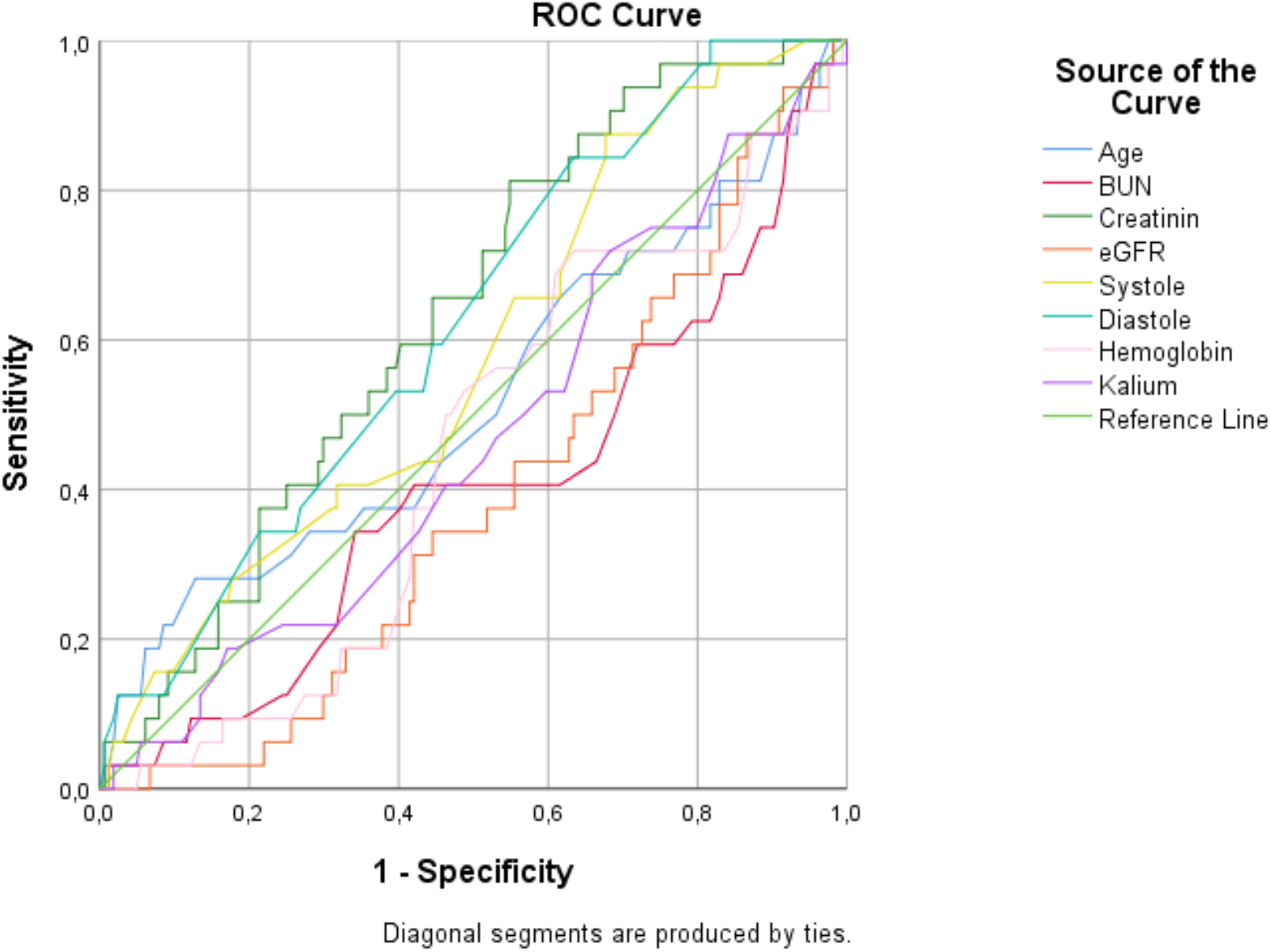
Receiver operator characteristic analysis of the clinical prediction model for age, BUN, serum creatinine, eGFR, systolic blood pressure, diastolic blood pressure, hemoglobin level and serum potassium. Prediction of LVH by age, BUN, serum creatinine, eGFR, systolic blood pressure, diastolic blood pressure, hemoglobin level and serum potassium. *P* < 0.001 for all ROC curves compared with 0.5 curves.

### Cut-off value for serum creatinine and other biomarkers to discriminate peaked T wave

The cut-off value of serum creatinine to discriminate peaked T wave can be seen in ROC Curve (Figure 4). For bivariate analysis, serum creatinine is positively discriminate peaked T wave based on ROC curve with area under curve (AUC) 0.708 (*P*<0.001) with the best cut off serum creatinine level at 9.03 (sensitivity 77% and specificity 61%) (Table 8).

**Table 8.** Area under the curve (AUC) of clinical prediction model for age, BUN, serum creatinine, eGFR, systolic blood pressure, diastolic blood pressure, hemoglobin level and serum potassium. Prediction of peaked T wave by age, BUN, serum creatinine, eGFR, systolic blood pressure, diastolic blood pressure, hemoglobin level and serum potassium. *P* < 0.001 for all ROC curves compared with 0.5 curves.

| Area Under the Curve |  |  |  |  |  |
| --- | --- | --- | --- | --- | --- |
| Test Result Variable(s) | Area | Std. Error<sup>a</sup> | Asymptotic Sig.<sup>b</sup> | Asymptotic 95% Confidence Interval |  |
|  |  |  |  | Lower Bound | Upper Bound |
| Age | ,564 | ,048 | ,201 | ,469 | ,658 |
| BUN | ,430 | ,048 | ,163 | ,337 | ,524 |
| Creatinin | ,707 | ,047 | ,000 | ,616 | ,798 |
| eGFR | ,303 | ,048 | ,000 | ,209 | ,398 |
| Systole | ,545 | ,048 | ,370 | ,451 | ,639 |
| Diastole | ,566 | ,049 | ,189 | ,469 | ,662 |
| Hemoglobin | ,447 | ,050 | ,286 | ,348 | ,545 |
| Kalium | ,688 | ,044 | ,000 | ,601 | ,775 |

The test result variable(s): Age, BUN, Creatinin, Systole, Diastole, Hemoglobin, Kalium has at least one tie between the positive actual state group and the negative actual state group. Statistics may be biased.
a. Under the nonparametric assumption
b. Null hypothesis: true area = 0.5

**Figure 4.**
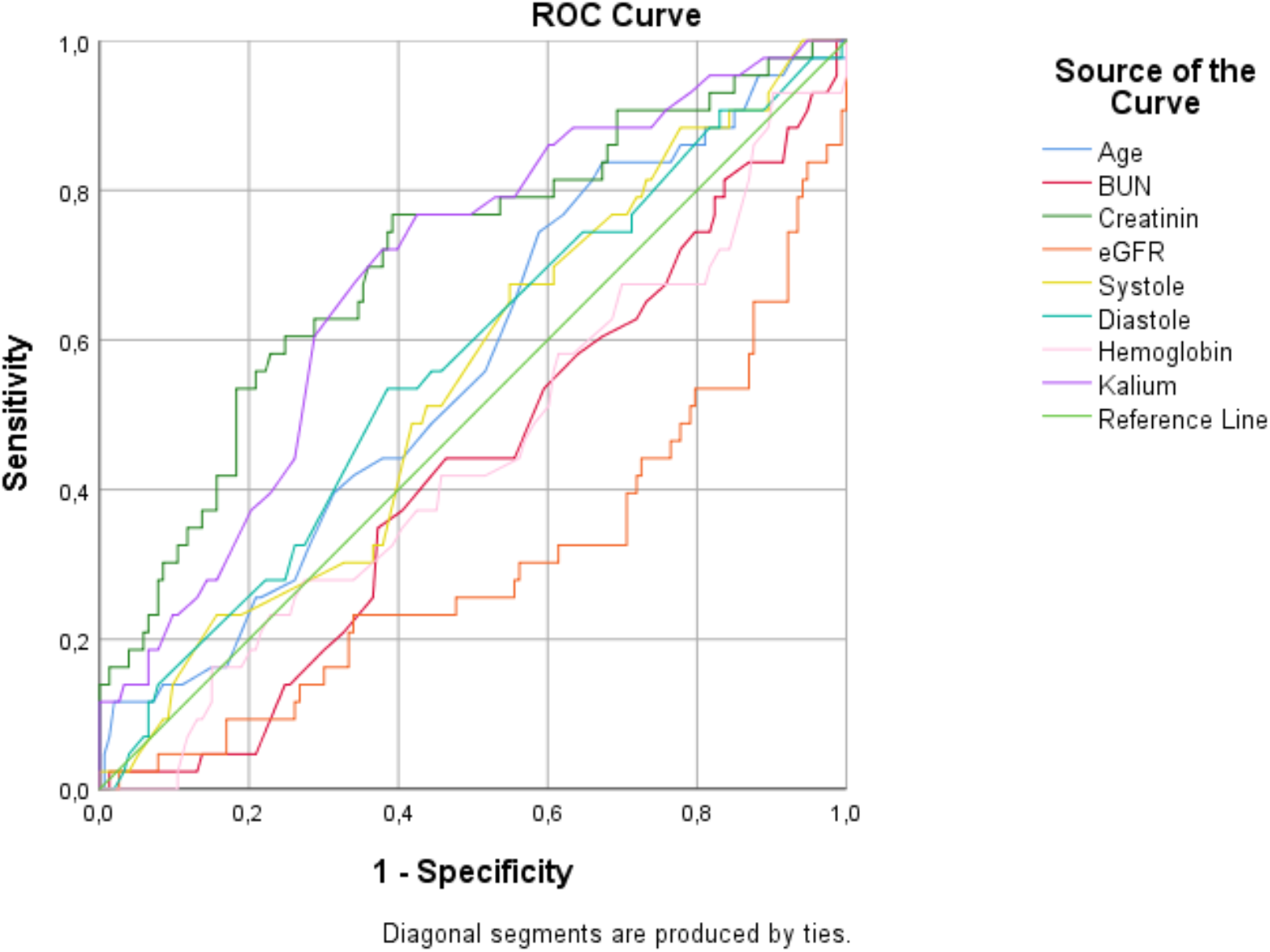
Receiver operator characteristic analysis of the clinical prediction model for age, BUN, serum creatinine, eGFR, systolic blood pressure, diastolic blood pressure, hemoglobin level and serum potassium. Prediction of peaked T wave by age, BUN, serum creatinine, eGFR, systolic blood pressure, diastolic blood pressure, hemoglobin level and serum potassium. *P* < 0.001 for all ROC curves compared with 0.5 curves.

### Cut-off value for hemoglobin concentration and other biomarkers to discriminate poor precordial R wave progression

For bivariate analysis, serum creatinine was a negative predictor for poor precordial R wave progression based on ROC curve (Figure 5) with AUC 0.464 (Table 9).

**Table 9.** Area under the curve (AUC) of clinical prediction model for age, BUN, serum creatinine, eGFR, systolic blood pressure, diastolic blood pressure, hemoglobin level and serum potassium. Prediction of poor precordial R wave progression by age, BUN, serum creatinine, eGFR, systolic blood pressure, diastolic blood pressure, hemoglobin level and serum potassium. *P* < 0.001 for all ROC curves compared with 0.5 curves.

| Test Result Variable(s) | Area Under the Curve |  |  |  |  |
| --- | --- | --- | --- | --- | --- |
|  | Area | Std. Error<sup>a</sup> | Asymptotic Sig.<sup>b</sup> | Asymptotic 95% Confidence Interval |  |
|  |  |  |  | Lower Bound | Upper Bound |
| Age | ,431 | ,047 | ,148 | ,338 | ,523 |
| BUN | ,497 | ,048 | ,957 | ,404 | ,591 |
| Creatinin | ,464 | ,045 | ,449 | ,376 | ,551 |
| eGFR | ,517 | ,046 | ,725 | ,427 | ,606 |
| Systole | ,523 | ,050 | ,629 | ,426 | ,621 |
| Diastole | ,479 | ,048 | ,655 | ,385 | ,572 |
| Hemoglobin | ,532 | ,047 | ,504 | ,439 | ,625 |
| Kalium | ,491 | ,051 | ,846 | ,391 | ,590 |

The test result variable(s): Age, BUN, Creatinin, Systole, Diastole, Hemoglobin, Kalium has at least one tie between the positive actual state group and the negative actual state group. Statistics may be biased.
a. Under the nonparametric assumption
b. Null hypothesis: true area = 0.5

**Figure 5.**
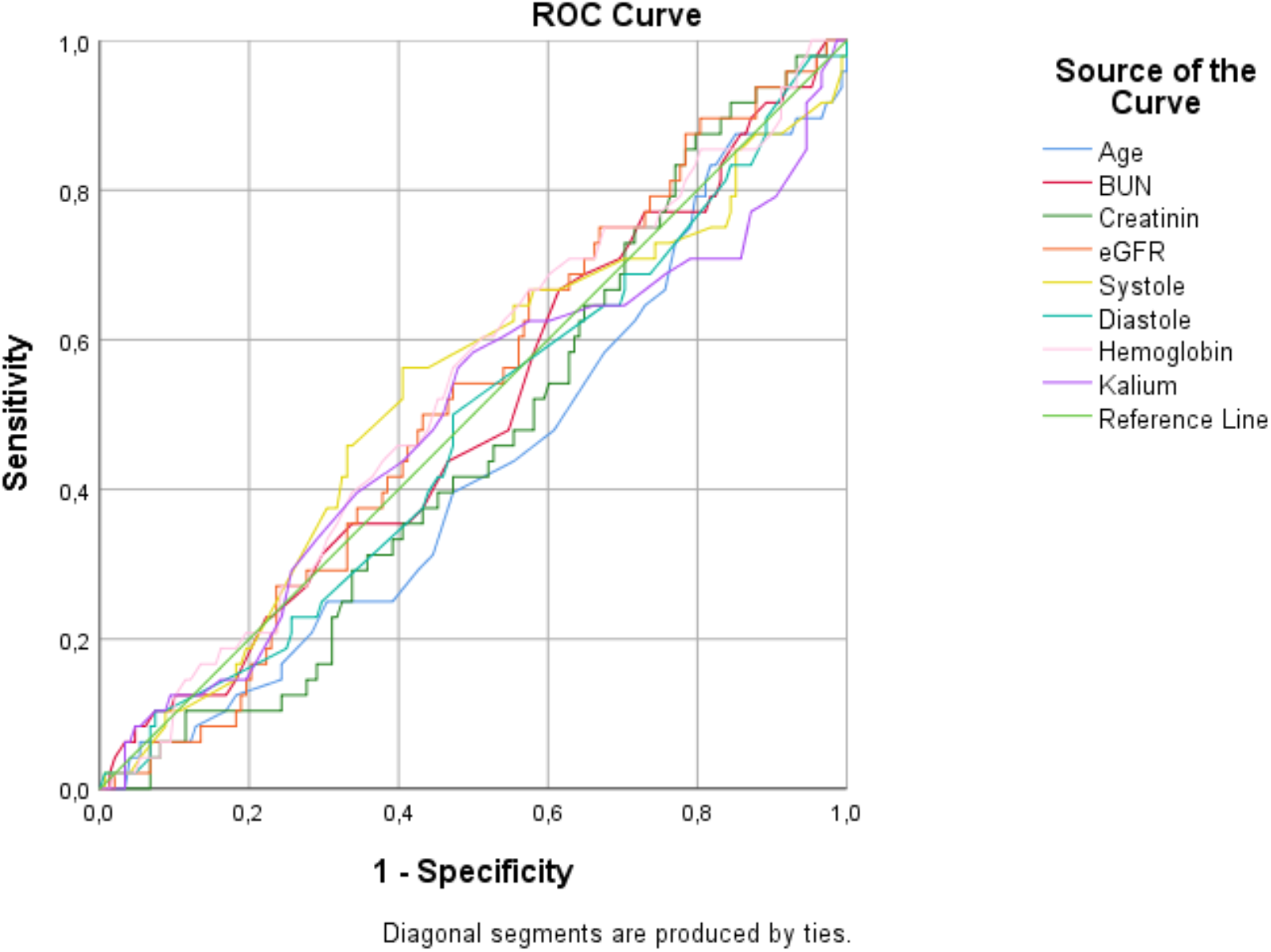
Receiver operator characteristic analysis of the clinical prediction model for age, BUN, serum creatinine, eGFR, systolic blood pressure, diastolic blood pressure, hemoglobin level and serum potassium. Prediction of poor precordial R wave progression by age, BUN, serum creatinine, eGFR, systolic blood pressure, diastolic blood pressure, hemoglobin level and serum potassium. *P* < 0.001 for all ROC curves compared with 0.5 curves.

### Cut-off value for hemoglobin concentration and other biomarkers to discriminate pathological Q wave

For bivariate analysis, serum creatinine was a negative predictor for pathological Q wave based on ROC curve (Figure 6) with AUC 0.397 (Table 10).

**Table 10.** Area under the curve (AUC) of clinical prediction model for age, BUN, serum creatinine, eGFR, systolic blood pressure, diastolic blood pressure, hemoglobin level and serum potassium. Prediction of pathological Q wave by age, BUN, serum creatinine, eGFR, systolic blood pressure, diastolic blood pressure, hemoglobin level and serum potassium. *P* < 0.001 for all ROC curves compared with 0.5 curves.

| Test Result Variable(s) | Area Under the Curve |  |  |  |  |
| --- | --- | --- | --- | --- | --- |
|  | Area | Std. Error<sup>a</sup> | Asymptotic Sig.<sup>b</sup> | Asymptotic 95% Confidence Interval |  |
|  |  |  |  | Lower Bound | Upper Bound |
| Age | ,389 | ,058 | ,060 | ,276 | ,502 |
| BUN | ,567 | ,055 | ,260 | ,459 | ,674 |
| Creatinin | ,397 | ,052 | ,082 | ,295 | ,499 |
| eGFR | ,595 | ,053 | ,106 | ,492 | ,699 |
| Systole | ,526 | ,059 | ,655 | ,411 | ,641 |
| Diastole | ,516 | ,055 | ,783 | ,408 | ,624 |
| Hemoglobin | ,641 | ,061 | ,017 | ,521 | ,760 |
| Kalium | ,482 | ,063 | ,758 | ,359 | ,605 |

The test result variable(s): Age, BUN, Creatinin, Systole, Diastole, Hemoglobin, Kalium has at least one tie between the positive actual state group and the negative actual state group. Statistics may be biased.
a. Under the nonparametric assumption
b. Null hypothesis: true area = 0.5

**Figure 6.**
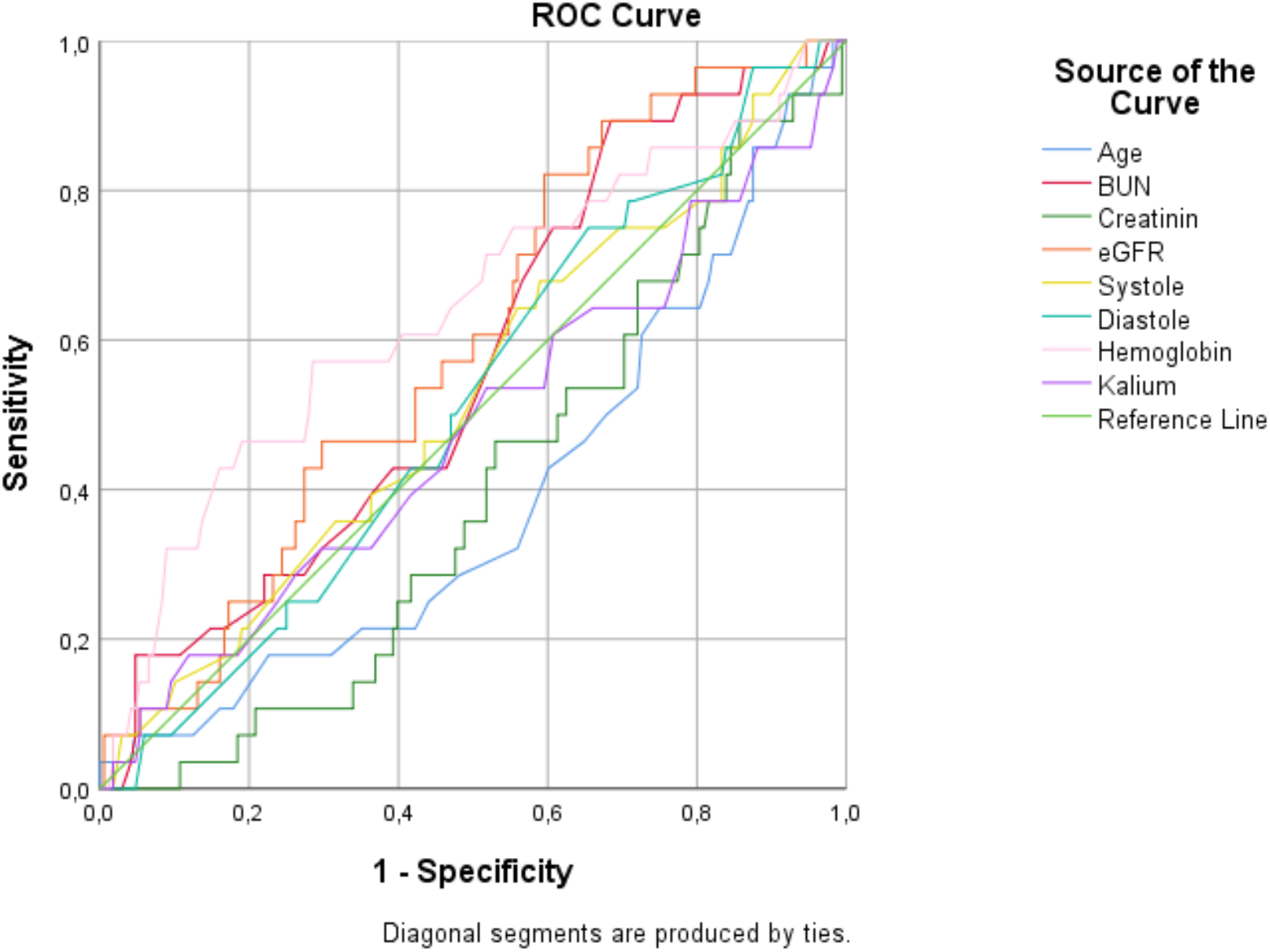
Receiver operator characteristic analysis of the clinical prediction model for age, BUN, serum creatinine, eGFR, systolic blood pressure, diastolic blood pressure, hemoglobin level and serum potassium. Prediction of pathological Q wave by age, BUN, serum creatinine, eGFR, systolic blood pressure, diastolic blood pressure, hemoglobin level and serum potassium. *P* < 0.001 for all ROC curves compared with 0.5 curves.

### Cut-off value for hemoglobin concentration and other biomarkers to discriminate non-specific ST-T changes

For bivariate analysis, serum creatinine was a negative predictor for non-specific ST-T changes based on ROC curve (Figure 7) with AUC 0.406 (Table 11).

**Table 11.** Area under the curve (AUC) of clinical prediction model for age, BUN, serum creatinine, eGFR, systolic blood pressure, diastolic blood pressure, hemoglobin level and serum potassium. Prediction of non-specific ST-T changes by age, BUN, serum creatinine, eGFR, systolic blood pressure, diastolic blood pressure, hemoglobin level and serum potassium. *P* < 0.001 for all ROC curves compared with 0.5 curves.

| Test Result Variable(s) | Area Under the Curve |  |  |  |  |
| --- | --- | --- | --- | --- | --- |
|  | Area | Std. Error<sup>a</sup> | Asymptotic Sig.<sup>b</sup> | Asymptotic 95% Confidence Interval |  |
|  |  |  |  | Lower Bound | Upper Bound |
| Age | ,422 | ,061 | ,199 | ,303 | ,541 |
| BUN | ,485 | ,066 | ,811 | ,356 | ,614 |
| Creatinin | ,406 | ,058 | ,123 | ,292 | ,521 |
| eGFR | ,594 | ,058 | ,123 | ,481 | ,707 |
| Systole | ,521 | ,060 | ,729 | ,403 | ,639 |
| Diastole | ,494 | ,059 | ,916 | ,377 | ,610 |
| Hemoglobin | ,556 | ,053 | ,362 | ,451 | ,660 |
| Kalium | ,468 | ,060 | ,603 | ,351 | ,586 |

The test result variable(s): Age, BUN, Creatinin, Systole, Diastole, Hemoglobin, Kalium has at least one tie between the positive actual state group and the negative actual state group. Statistics may be biased.
a. Under the nonparametric assumption
b. Null hypothesis: true area = 0.5

**Figure 7.**
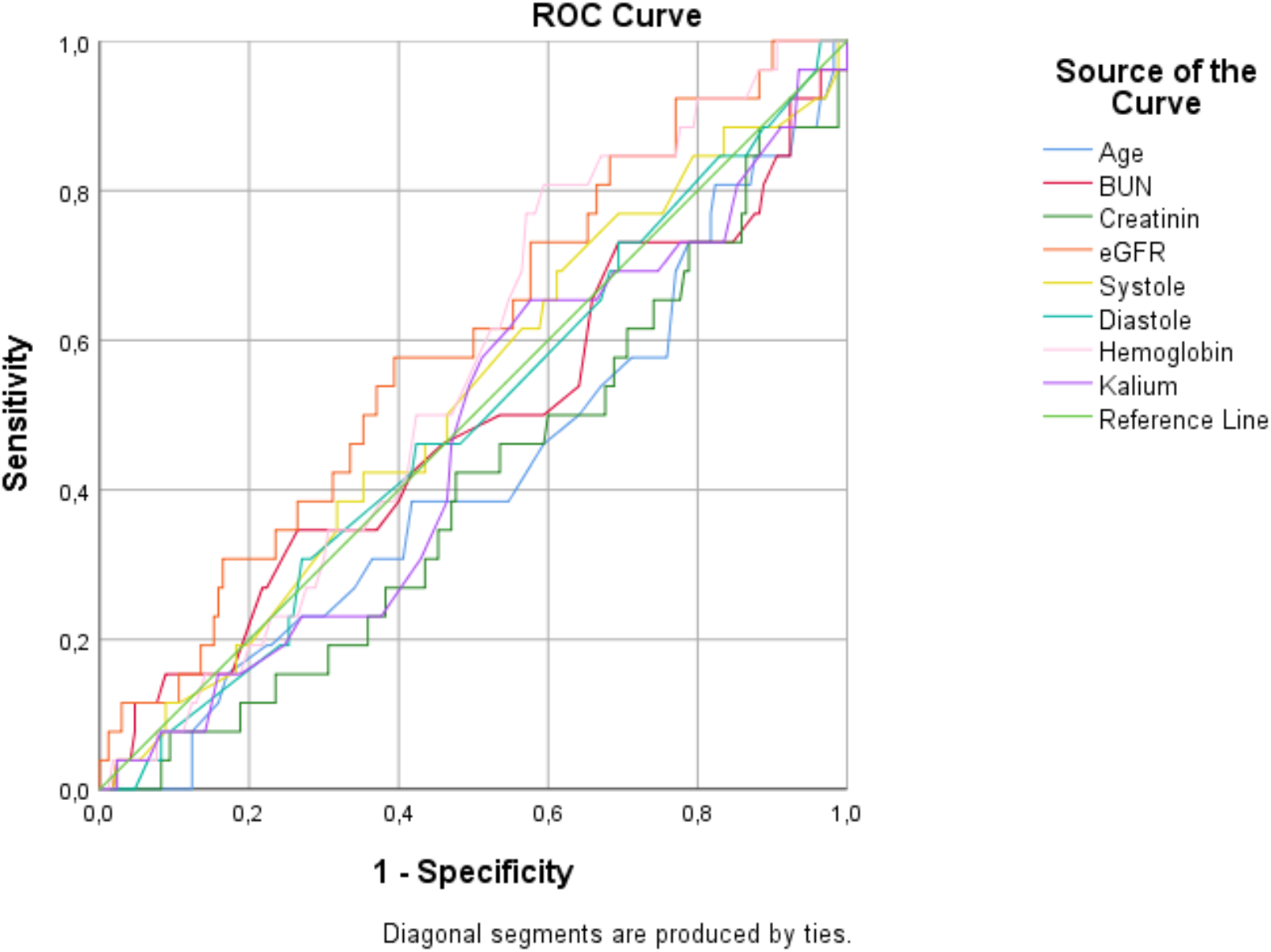
Receiver operator characteristic analysis of the clinical prediction model for age, BUN, serum creatinine, eGFR, systolic blood pressure, diastolic blood pressure, hemoglobin level and serum potassium. Prediction of non-specific ST-T changes by age, BUN, serum creatinine, eGFR, systolic blood pressure, diastolic blood pressure, hemoglobin level and serum potassium. *P* < 0.001 for all ROC curves compared with 0.5 curves.

### Cut-off value for hemoglobin concentration and other biomarkers to discriminate frontal axis deviation

For bivariate analysis, serum creatinine was a negative predictor for frontal axis deviation based on ROC curve (Figure 8) with AUC 0.462 (Table 12).

**Table 12.** Area under the curve (AUC) of clinical prediction model for age, BUN, serum creatinine, eGFR, systolic blood pressure, diastolic blood pressure, hemoglobin level and serum potassium. Prediction of frontal axis deviation by age, BUN, serum creatinine, eGFR, systolic blood pressure, diastolic blood pressure, hemoglobin level and serum potassium. *P* < 0.001 for all ROC curves compared with 0.5 curves.

| Test Result Variable(s) | Area Under the Curve |  |  |  |  |
| --- | --- | --- | --- | --- | --- |
|  | Area | Std. Error<sup>a</sup> | Asymptotic Sig.<sup>b</sup> | Asymptotic 95% Confidence Interval |  |
|  |  |  |  | Lower Bound | Upper Bound |
| Age | ,442 | ,067 | ,383 | ,311 | ,573 |
| BUN | ,526 | ,071 | ,697 | ,388 | ,664 |
| Creatinin | ,462 | ,065 | ,567 | ,335 | ,589 |
| eGFR | ,524 | ,067 | ,722 | ,393 | ,654 |
| Systole | ,443 | ,069 | ,391 | ,308 | ,578 |
| Diastole | ,477 | ,070 | ,729 | ,339 | ,615 |
| Hemoglobin | ,693 | ,067 | ,004 | ,561 | ,825 |
| Kalium | ,467 | ,062 | ,627 | ,346 | ,589 |

The test result variable(s): Age, BUN, Systole, Diastole, Hemoglobin, Kalium has at least one tie between the positive actual state group and the negative actual state group. Statistics may be biased.
a. Under the nonparametric assumption
b. Null hypothesis: true area = 0.5

**Figure 8.**
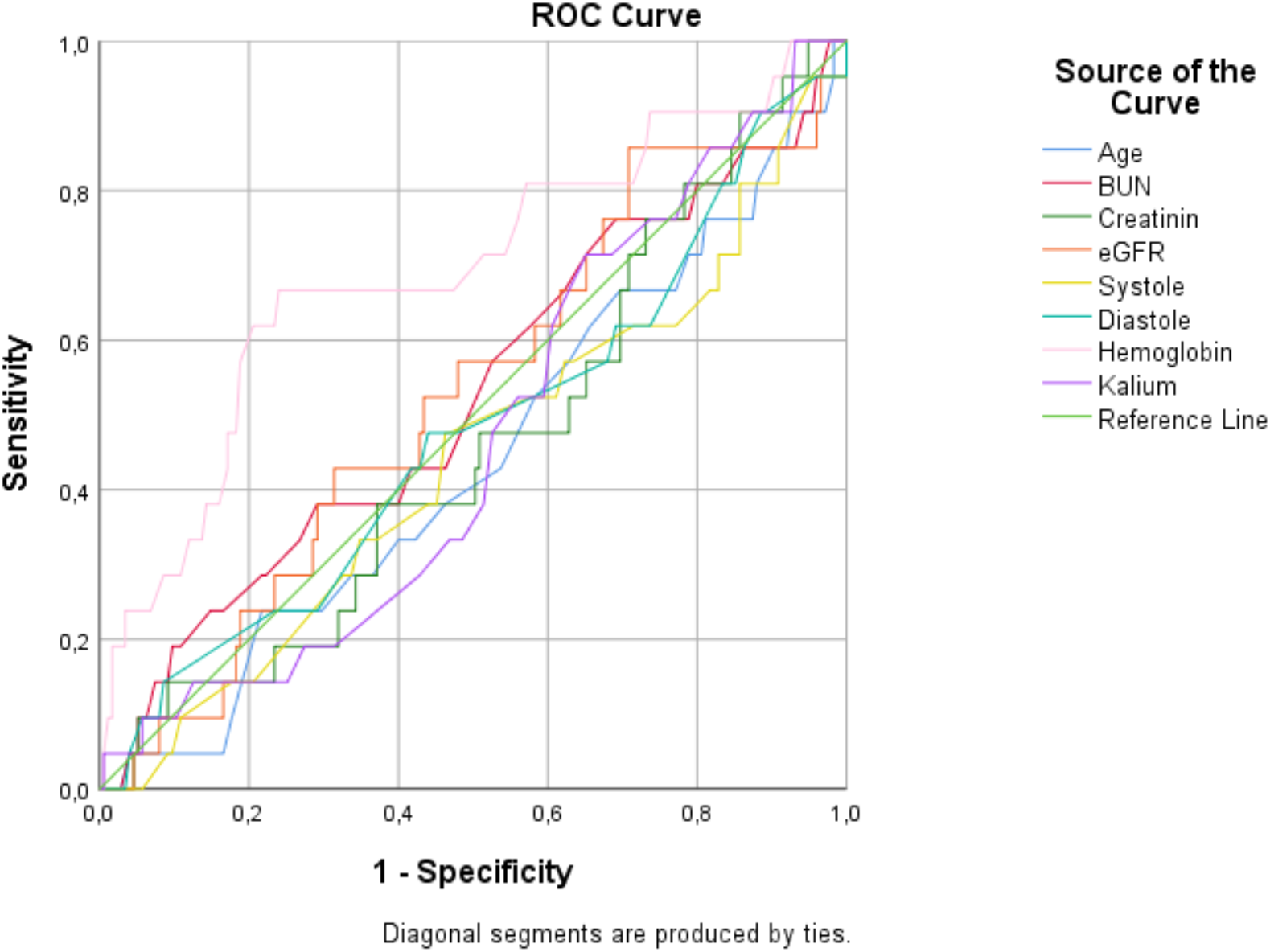
Receiver operator characteristic analysis of the clinical prediction model for age, BUN, serum creatinine, eGFR, systolic blood pressure, diastolic blood pressure, hemoglobin level and serum potassium. Prediction of frontal axis deviation by age, BUN, serum creatinine, eGFR, systolic blood pressure, diastolic blood pressure, hemoglobin level and serum potassium. *P* < 0.001 for all ROC curves compared with 0.5 curves.

### Univariate and multivariate analysis for ECG abnormalities

A multivariate analysis was used to see the most important factor for ECG abnormalities in CKD patients. Prolong QTc interval and fragmented QRS was used as the marker of ECG abnormality, because of their importance as predictors of long-term mortality and also as the most common abnormal ECG findings in our population. There were 110 (55.55%) patients who have QTc interval prolongation or fragmented QRS, or both. The variables whose *P*-value <0.25 in univariate analysis from baseline characteristics and laboratory findings were specified as potential risk markers and included in the full model (Table 13). In the univariate analysis, blood urea nitrogen (BUN), systolic and diastolic blood pressure, and potassium level were identified as potential risk predictors. However, in multivariate logistic regression analysis, only higher systolic blood pressure was determined as an independent predictor of abnormal ECG finding in CKD patients, as systolic blood pressure increase by one unit, the odds of having abnormal ECG are increased 1.02 times (95% CI: 1.00 – 1.02, *p*=0.042). Further analysis was done using ROC curve to determine systolic blood pressure cut off, and the best optimal cut off was 140 mmHg (sensitivity 60%, specificity 59%, AUC 0.57).

**Table 13.** Univariate and multivariate logistic regression for prediction of ECG abnormalities among chronic kidney disease patients

| Variable | Univariate Analysis |  |  | Multivariate Analysis |  |  |
| --- | --- | --- | --- | --- | --- | --- |
|  | HR | 95% CI | *P* value | HR | 95% CI | *P* value |
| Age | 0.993 | 0.970 - 1.017 | 0.568 |  |  |  |
| Man gender | 0.991 | 0.566 - 1.736 | 0.975 |  |  |  |
| BUN | 0.983 | 0.959 - 1.007 | 0.164 |  |  |  |
| Creatinine | 1.009 | 0.974 – 1.045 | 0.621 |  |  |  |
| eGFR | 0.997 | 0.978 - 1.017 | 0.793 |  |  |  |
| Systolic blood pressure | 1.011 | 1.001 - 1.022 | 0.038 | 1.018 | 1.000 – 1.021 | 0.042 |
| Diastolic blood pressure | 1.011 | 0.994 - 1.029 | 0.215 |  |  |  |
| Kalium | 0.829 | 0.634 - 1.084 | 0.170 |  |  |  |

## DISCUSSION

The long-term CKD influences the pathogenesis of cardiorenal syndromes, cardiovascular diseases, cardiovascular risk factors, and substantial cardiovascular mortality, there is a need to screen Indonesian CKD patients who are at risk of getting earlier complications [9]. Renal dysfunction is an independent and significant contributor to worse cardiovascular outcomes[10]. Serum creatinine levels are frequently used as a screening test to assess renal dysfunction[11]. This study also explored the potential role of serum creatinine as a parameter for hospitalized CKD patients for getting cardiovascular diseases and/or cardiac arrhythmias. Even though serum creatinine is conventional and old-fashioned diagnostic tools, however, it is cost-effective, rapidly, and highly applicable to larger cohorts of CKD patients compared with the more expensive, more sophisticated, more laborious, time-consuming serum human neutrophil gelatinase-associated interleukin-18 (IL-18), cystatin-C (cys-C), lipocalin (NGAL), kidney injury molecule-1 (KIM-1), fibroblast growth factor-23 (FGF-23), carboxy-terminal fragment of insulin-like growth factor binding protein-4 (CT-IGFBP-4) and liver-type fatty acid-binding proteins (L-FABP) required for the identification of early cardiovascular diseases in acute kidney injury patients[12, 13].

In our model, serum creatinine had the highest AUC value to predict left ventricular hypertrophy (LVH). Our results might be supported by Leoncini et al (2004), which concluded that creatinine clearance proved to be a very sensitive marker of clinical and subclinical cardiovascular damage[14]. LVH is a well-established sign of cardiovascular damage and also an independent predictor of future cardiovascular morbidity in CKD patients[14]. Although conventional ECG has been thought to be a less specific method than echocardiography for detecting LVH, however, its specificity is even greater than echocardiography. Thus, the more severe the LVH is on echocardiography, the more likely it would be diagnosed on ECG[15]. Serum creatinine also had higher AUC to predict peaked T wave and long QTc interval, compared to serum potassium. This fact might be interesting because serum potassium had long been used as a strong predictor for peaked T wave. Activation of the potassium channels can stimulate speeding up membrane repolarisation, cause inactivates sodium channels can make membrane become depolarized[16]. in hyperkalemia, typical ECG findings progress from shortened QTc interval, and tall, peaked T waves [17]. Hyperkalemia stimulates activation of potassium channel and inactivation of sodium channels which causes shortening QTc interval, sluggish cardiac conduction, widening of the QRS complex and smaller P waves [16]. Moreover, in our model, serum creatinine showed a stronger predictor for peaked T wave compared to serum potassium in hospitalized CKD patients. This fact might be explained by an absence of hydro-electrolytic alterations in many CKD patients. However, calcium also plays a role in T wave amplitude. [18]. Another literature had published that hyperkalemia doesn’t always include changes in the abnormality ECG, especially in the amplitude of T wave, as a progressive elevation of serum potassium doesn’t always make the auricular muscle unresponsive[19, 20].

The phenomenon of higher serum creatinine correlate with long QTc interval could be explained by numerous factors influencing the QT interval: 1) Patients with CKD often take many medications that can cause QT prolongation; 2) Secondary hyperparathyroidism, a common complication of CKD, could lead to hypocalcemia and hypomagnesemia; 3) Renal anemia, macrocytosis and anisocytosis are also related to prolonged QT intervals due to subsequent hypoxia, autonomic dysfunction and decreased myocardial oxygen supply; 4) Impairment of delayed rectifier potassium channels; 5) Hypertension can significantly reduce potassium current densities (I_peak_, I_to_, I_Kur_, I_ss_, and I_k1_) and increase the L-type calcium channel; and 6) Reactive oxygen species and uremic toxin leading to inflammatory condition and oxidative stress itself may also predispose to cardiac electrical remodeling[21, 22]. Therefore, the prolongation of the QTc interval is common in hospitalized patients with CKD.

Hemoglobin concentration often reflects duration and chronicity in individuals with CKD, although its level fluctuates frequently [23]. Recent studies suggest that a lower hemoglobin concentration is related with subsequent cardiovascular mortality and increased risk of coronary artery disease in hospitalized patients with CKD, such as CONFIRM-HF Trial [24], IRON OUT HF Trial [25], TIME Trial [26], GUSTO IIb Trial [27], PURSUIT Trial [28], PARAGON B Trial [29], ANCHOR Study [30], and ARIC Study [31]. On the other hand, our study explained that higher hemoglobin concentration showed a strong predictor for pathological Q wave, non-specific ST-T changes, and frontal axis deviation. We believed that the effect of higher hemoglobin concentrations on ECG abnormalities varies across various subtypes of cardiovascular diseases. If the state of anemia continues chronically, the hemodynamic changes found in anemia can contribute to left ventricular hypertrophy and progressive arterial walls [32]. In addition, the hemodynamic changes found in the higher hemoglobin concentration might be primarily determined by the viscosity of blood. Greater hematocrit concentrations would thus significantly slowing its flow rate throughout the body, thicken the blood, raising the peripheral vascular resistance, and reducing coronary blood flow and perfusion to various tissues including the cardiomyocyte. The linked, mechanisms, pathogenesis and pathophysiology of coronary artery disease (CAD) in CKD are complex. Many shreds of evidence clearly stated a significant association between lower or higher hemoglobin concentration and the development of CAD or poor outcomes in patients with known CAD, Therefore, ECG abnormalities such as pathological Q wave (reflects previous myocardial infarction), non-specific ST/T changes (reflects intrinsic myocardial diseases i.e. ischemia) and frontal axis deviation (sign of hypertrophy or reduced muscle mass or conduction abnormalities) are just suggesting the existence of the relationship between CKD and cardiovascular complication.

Because hypertension in CKD is predominantly systolic [33], we explored the relationship between systolic blood pressure (BP) with ECG abnormalities in a cross-sectional study. In our logistic regression models, only systolic BP is associated with ECG abnormalities (QTc interval prolongation or fragmented QRS, or both). The severity of diastolic dysfunction in CKD usually progressed (from grade I to grade III) with the rise in systolic BP readings. The presence of diastolic dysfunction itself also the risk factor to develop any ECG abnormalities. We did not find an association between diastolic BP with ECG abnormalities, even in bivariate analysis, regardless of BP technique. One possible reason for the lack of association between diastolic BP with ECG abnormalities maybe because the mean diastolic BP in our study population is nearly normal. In perspective, the differences between predictors of systolic BP and diastolic BP might be explainable [34].

Our study suffers from many limitations. Firstly, further risk factors for ECG abnormalities were not taken into account for the analysis. Secondly, this was a cross-sectional study, single-center with small sample sizes in once measurement and therefore we cannot imply causality from association studies. Thirdly, necessary information such as echocardiography or chest x-ray may be unavailable. Fourthly, we did not have data on medications that may have influenced cardiovascular parameters in this study. Fifthly, the interpretation of several laboratories and clinical parameters changes over time. For example, serum creatinine is a breakdown product of muscle, its level is directly associated with muscle mass, which is lower in the elderly, women and whites[35]. The strength of this study was that data already exists, prevalence estimation could be done at one point in time, complete study populations minimizing selection bias and independently collected data.

## CONCLUSION

In hospitalized CKD patients, ECG abnormalities are common, where fragmented QRS and prolonged QTc were the most prevalent ECG abnormalities. Serum creatinine and hemoglobin could predict peaked T wave and prolonged QTc among hospitalized CKD patients. Systolic blood pressure could predict prolonged QTc and fragmented QRS in CKD patients. Longitudinal studies are needed to show the causality of each parameter with ECG abnormalities.

## Data Availability

The datasets generated during and/or analyzed during the current study are not publicly available due to protecting participant confidentiality but are available from the corresponding author on reasonable request.

## Ethical Clearance

Dr. Soetomo General Academic Hospital conferred ethical clearance for this study. Compliance with ethical standards, this research proposal was approved by the ethical committees of Dr. Soetomo General Academic Hospital in collaboration with Universitas Airlangga College of Medicine Research Ethics Council (Ref: 1811/KEPK/II/2020) under the name of Rerdin Julario as the Principal Investigator.

## Acknowledgment

The authors would also like to offer special thanks to all staff, fellows, residents, and nurses from the Department of Cardiology and Vascular Medicine & Divison of Nephrology Department of Medicine, Dr. Soetomo Academic General Hospital, Surabaya for their technical contribution.

## Data Availability Statements

The datasets generated and analyzed during this study are not publicly available but are available by contacting the corresponding author.

## Author Contribution

R.J. conceived the idea. R.A.N. designed the research and initially wrote the manuscript. B.P.D.K. and T.S.P. carried out the observations. E.P.B.M. helped the project administration. R.E.I. analyzed and interpreted the data, developed the model formulation. F.F.A. helped draft and reviewed the manuscript. All authors read and approved the final version of the manuscript.

## Provenance and peer review

Not commissioned; externally peer reviewed.

